# Smoking is Associated with COVID-19 Progression: A Meta-Analysis

**DOI:** 10.1101/2020.04.13.20063669

**Authors:** Roengrudee Patanavanich, Stanton A. Glantz

**Author notes:** Corresponding author: Stanton Glantz, PhD, Center for Tobacco Control Research and Education, 530 Parnassus Avenue, Suite 366, University of California, San Francisco, San Francisco, CA, U.S.A. 94143-1390.

## Abstract

**Objective:** To determine the association between smoking and progression of COVID-19.

**Design:** A meta-analysis of 12 published papers.

**Data Source:** PubMed database was searched on April 6, 2020.

**Eligibility criteria and data analysis:** We included studies reporting smoking behavior of COVID-19 patients and progression of disease. Search terms included “smoking”, “smoker*”, “characteristics”, “risk factors”, “outcomes”, and “COVID-19”, “COVID”, “coronavirus”, “sar cov-2”, “sar cov 2”. There were no language limitations. One author extracted information for each study, screened the abstract or the full text, with questions resolved through discussion among both authors. A random effects meta-analysis was applied.

**Main Outcome Measures:** The study outcome was progression of COVID-19 among people who already had the disease.

**Results:** We identified 12 papers with a total of 9,025 COVID-19 patients, 878 (9.7%) with severe disease and 495 with a history of smoking (5.5%). The meta-analysis showed a significant association between smoking and progression of COVID-19 (OR 2.25, 95% CI 1.49-3.39, *p*=0.001). Limitations in the 12 papers suggest that the actual risk of smoking may be higher.

**Conclusions:** Smoking is a risk factor for progression of COVID-19, with smokers having higher odds of COVID-19 progression than never smokers. Physicians and public health professionals should collect data on smoking as part of clinical management and add smoking cessation to the list of practices to blunt the COVID-19 pandemic.

**What is already known on this topic:**

- Smoking increases risk and severity of pulmonary infections because of damage to upper airways and a decrease in pulmonary immune function.

**What this study adds:**

- Smoking is associated with COVID-19 severity.
- Smoking history should be part of clinical management of COVID-19 patients and cessation should be added to the list of practices to blunt the COVID-19 pandemic.

## Introduction

COVID-19, the coronavirus-transmitted infectious disease, has caused a worldwide pandemic. Smoking^1 2^ and e-cigarette use^3^ increase risk and severity of pulmonary infections because of damage to upper airways and a decrease in pulmonary immune function. In particular, smokers have a higher risk of infection and mortality from Cov-MERS.^4^ Two reviews^5 6^ of the first 5 papers presenting data on smoking and COVID-19 reached different conclusions. Another review described 6 published case series presenting data on smoking among COVID-19 patients but did not draw a conclusion about the association of severity of COVID-19 with smoking.^7^ We reviewed and summarized 12 papers presenting data on the association between smoking and severity of COVID-19.

## Methods

We conducted a systematic search using PubMed database on April 6, 2020, with the search term: ((smoking) OR (characteristics) OR (risk factors) OR (outcomes) OR (smoker*)) AND ((COVID-19) OR (COVID) OR (coronavirus) OR (sars cov-2) OR (sars cov 2)) for studies published between January 1, 2020 and April 6, 2020. There were no language restrictions. A total of 396 studies were retrieved through the search, of which 12, 10 from China,^8-17^ 1 from Korea,^18^ and 1 from the US,^19^ included data on smoking behavior and COVID-19 disease progression (Table 1). Ten studies^8 10-18^ were based on hospitalized patients and two^9 19^ included both hospitalized patients and outpatients.

**Table 1.** Characteristics of studies included in the analysis.

| Author | Study design | Time of data collection | Setting | Population | Definition of Disease Progression | Smoking status | Smoking prevalence |
| --- | --- | --- | --- | --- | --- | --- | --- |
| CDC COVID-19 Response Team<sup>19</sup> (US) | Retrospective | 12 Feb 2020-28 Mar 2020 | United States | 7,162 confirmed COVID-19 patients with completed information* | ICU admission | Current, former, never smoker | 3.6% |
| Dong X, et al.<sup>8</sup> | Case series | 20 Jan 2020-3 Mar 2020 | Zhongnan hospital of Wuhan University, No. 7 hospital of Wuhan, and Wuhan children's hospital, China | 9 adults hospitalized patients with laboratory confirmed COVID-19** | Bilateral pneumonia, respiratory distress, required mechanical ventilation, ICU care, or hospitalized > 10 days. | Current smoker | 11.1% |
| Guan WJ, et al.<sup>9</sup> | Retrospective | 11 Dec 2019-29 Jan 2020 | 552 hospitals in 30 provinces, autonomous regions, and municipalities in China | 1,099 patients (both hospitalized and outpatient) with laboratory confirmed COVID-19 | ICU admission, the use of mechanical ventilation, or death | Current, former, never smoker | 14.6% |
| Huang C, et al.<sup>10</sup> | Retrospective | 16 Dec 2019-2 Jan 2020 | Jin Yin-tan hospital in Wuhan, China | 41 hospitalized patients with laboratory confirmed COVID-19 | ICU admission | Current smoker | 7.3% |
| Kim ES, et al.<sup>18</sup> (Korea) | Retrospective | 19 Jan 2020-17 Feb 2020 | Korea | 28 confirmed COVID-10 cases nationwide. | Required oxygen supplement | Smoking | 18.5% |
| Liu W, et al.<sup>11</sup> | Retrospective | 30 Dec 2019-15 Jan 2020 | Three tertiary hospitals in Wuhan, China | 78 hospitalized patients with laboratory confirmed COVID-19 | Respiratory distress, respiratory failure, required mechanical ventilation and ICU care, or death | History of smoking | 6.4% |
| Mo P, et al.<sup>12</sup> | Retrospective | 1 Jan 2020-5 Feb 2020 | Zhongnan hospital of Wuhan University, China | 155 hospitalized patients with laboratory confirmed COVID-19 | Did not improve after treatment, status changed to severe, or hospitalized > 10 days | Current smoker | 3.9% |
| Shi Y, et al.<sup>13</sup> | Retrospective | Up to 17 Feb 2020 | Zhejiang province of China | 487 hospitalized patients with laboratory confirmed COVID-19 | Severe disease (without precisely defining it) or death | History of smoking | 8.4% |
| Wan S, et al.<sup>14</sup> | Retrospective | 23 Jan 2020-8 Feb 2020 | Chongqing University Three Gorges hospital, in northeast Chongqing, China | 135 hospitalized patients with laboratory confirmed COVID-19 | Respiratory distress with respiratory rate $\geq 30$/min, or oxygen saturation $\leq 93\%$ at rest, or oxygenation index $\leq 300$ mmHg | Current smoker | 6.7% |
| Yang X, et al.<sup>15</sup> | Retrospective | 24 Dec 2019-26 Jan 2020 | Wuhan Jin Yin-tan hospital, Wuhan, China | 52 critically ill hospitalized patients with laboratory confirmed COVID-19 | Death | History of smoking | 3.8% |
| Zhang JJ, et al.<sup>16</sup> | Retrospective | 16 Jan 2020-3 Feb 2020 | No.7 Hospital of Wuhan, China | 140 hospitalized patients with laboratory confirmed COVID-19 | Respiratory distress with respiratory rate $\geq 30$/min, or oxygen saturation $\leq 93\%$ at rest, or oxygenation index $\leq 300$ mmHg | Current, former, never smoker | 6.4% |
| Zhou F, et al.<sup>17</sup> | Retrospective | 29 Dec 2019-31 Jan 2020 | Jin Yin-tan hospital and Wuhan Pulmonary hospital in Wuhan, China | 191 hospitalized patients with laboratory confirmed COVID-19 | Death | Current smoker | 5.8% |
| * Hospitalization status unknown was excluded from the analysis.<br>** Two children were excluded from the analysis. |  |  |  |  |  |  |  |

The exposure group is those who had a history of smoking (current smokers or former smokers) and unexposed group was never smokers. Five studies^8 10 12 14 17^assessed whether the patient was a “current smoker,” three studies^9 16 19^ assessed whether the patient was a current or former smoker (as separate categories), and four studies^11 13 15 18^ assessed whether the patent had a “history of smoking” (current or former).

Outcome is progression of COVID-19 to more severe or critical conditions or death (Table 1). Two studies^14 16^ categorized the outcome as severe (respiratory distress with respiratory rate ≥ 30/min, or oxygen saturation ≤93% at rest, or oxygenation index ≤300 mmHg, based on the diagnostic and treatment guideline for SARS-CoV-2 issued by Chinese National Health Committee^16^) or mild, three^8 11 12^ categorized the outcome as progression or improvement, two^10 19^ categorized the outcome as ICU admission or non-ICU admission, one^9^ categorized the outcome as the primary composite end point (ICU admission, the use of mechanical ventilation, or death) or not, two^15 17^ categorized the outcome as death or survivor, one^13^ categorized the outcome as the occurrence of severe cases (without defining severe) or death or mild, and one^18^ categorized the outcome as clinical deterioration during the hospitalization and needed supplemental oxygen therapy.

We computed unadjusted odds ratios (OR) and 95% confidence interval (CI) for each study using the number of smokers (current and former) and never smokers with and without disease progression. Random effects meta-analysis was performed using the Stata 14.0 *metan* command and using *metabias* command with Harbord and Peters to test for the presence of publication bias.

Patients or the public were not involved in the design, or conduct, or reporting of our research. We will widely disseminate our findings working with the media and interested clinical and public health groups.

## Results

A total of 9,025 COVID-19 patients included in our meta-analysis, 878 of whom (9.7%) experienced disease progression and 495 with a history of smoking (5.5%). A total of 88 patients with a history of smoking (17.8%) experienced disease progression, compared with 9.3% of never smoking patients.

The meta-analysis showed an association between smoking and COVID-19 progression (OR 2.25, 95% CI 1.49-3.39, p=0.001) (Figure 1). There was not significant heterogeneity among the studies (I^2^=28.9%, *p*=0.162) or publication bias (Harbord’s *p*=0.155, Peters’ *p*=0.668).

**Figure 1.**
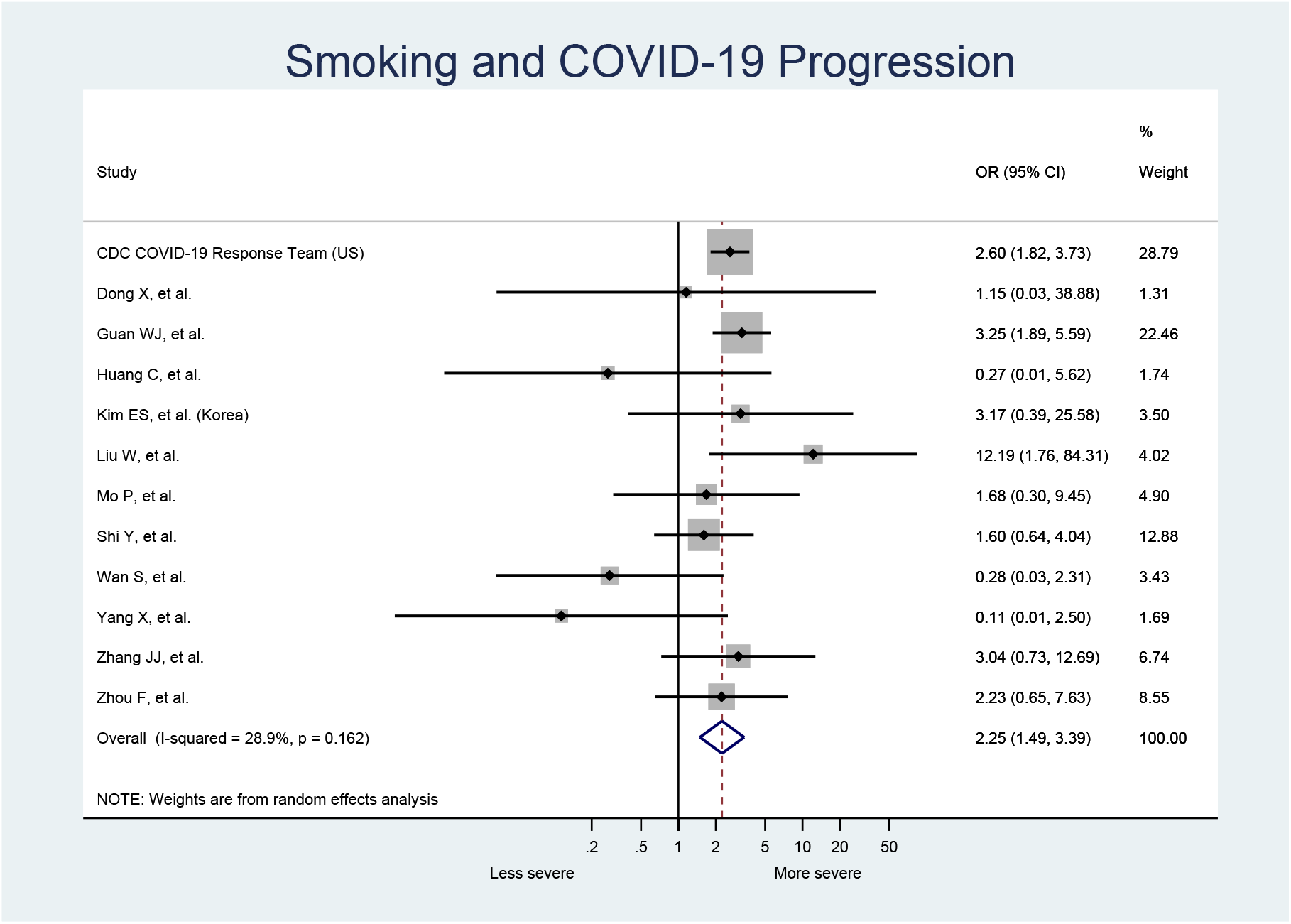
Smoking is associated with COVID-19 progression.

## Discussion

Our analysis confirms that smoking is a risk factor for progression of COVID-19, with smokers having 2.25-times the odds of severe COVID-19 outcomes than never smokers. This finding contrasts with an earlier meta-analysis,^6^ which included only 5 studies and used a non-standard method to compute the meta-analysis. The finding that smoking is associated with COVID-19 progression is not surprising because of the adverse effects of smoking on pulmonary immune function.^1 2^

Our study has several limitations.

The definition of “smoking” sometimes includes former smokers and sometimes does not. Only three studies^9 16 19^ separated current and former smokers in different categories, which was not enough data to do a meta-analysis for current and former smokers separately. Because the lung recovers after someone stops smoking, including former smokers in the exposed group biases the effect estimate to the null.

Reported smoking prevalence patients in all studies was substantially below smoking prevalence in the corresponding populations. Smoking prevalence in the 10 studies in China ranged from 3.8% to 14.6% vs 27.7% (52.1% for men and 2.7% for women) in the population in 2015,^20^ 18.5% in the Korean patients vs. 21.1%^21^ (37.0% for men and 5.2% for women) in 2017, and 3.6% in the US patients vs. 13.7%^22^ (15.6% for men and 12.0% for women) in 2018. It is highly likely that many smokers were misclassified as nonsmokers, which also biases the risk estimate toward the null.

We computed and assessed unadjusted odds ratios based on the numbers of patients reported in the studies. Only one^11^ of the studies reported unadjusted and adjusted ORs using multivariate analysis; after adjusting for confounding, the effect of smoking on disease severity was higher (unadjusted: OR 12.19, 95% CI 1.76-84.31, *p*=0.011; adjusted: OR 14.29, 95% CI 1.58-25.0, *p*=0.018).

None of these studies assessed e-cigarette use.

All these limitations suggest that this analysis underestimates the risk of smoking in terms of increasing COVID-19 severity.

All 12 studies were of patients who had already developed COVID-19, so the risk estimate we report does not represent the effect of smoking on the risk of contracting COVID-19 in the general population. As population-level testing ramps up, it would be useful to collect data on smoking and e-cigarette use to determine what risks these behaviors impose in terms of infection.

## Conclusions

Smoking is associated with COVID-19 disease progression. Physicians and public health professionals should collect data on smoking and, given the pulmonary effects of e-cigarettes,^3^ e-cigarette use, as part of routine clinical assessments and add smoking cessation to the list of practices to blunt the COVID-19 pandemic.

## Data Availability

All data are included in the manuscript.

## FUNDING

This work was supported by National Institute of Drug Abuse grant R01DA043950, cooperative agreement U54HL147127 from the National Heart, Lung, and Blood Institute and the Food and Drug Administration Center for Tobacco Products and the Faculty of Medicine Ramathibodi Hospital, Mahidol University, Thailand. The content is solely the responsibility of the authors and does not necessarily represent the official views of NIH or the Food and Drug Administration. The funding sources for this study had no role in the study design, data collection, data analysis, data interpretation, or the writing of the manuscript.

## COMPETITING INTERESTS

All authors have completed the Unified Competing Interest form (available on request from the corresponding author) and declare that the work was supported as described in the funding statement above. The authors have no financial relationships with any organisations that might have an interest in the submitted work in the previous three years and no other relationships or activities that could appear to have influenced the submitted work.

## CONTRIBUTORSHIP

RP developed the idea for the study, collected, analyzed the data, and wrote the first draft of the manuscript. SAG assisted with revising and refining the manuscript. Both authors affirm that the manuscript is an honest, accurate, and transparent account of the study being reported; that no important aspects of the study have been omitted; and that any discrepancies from the study as planned have been explained.

## ETHICAL APPROVAL

Not required.

## DATA SHARING STATEMENT

All data used to prepare this paper are available from the cited sources.

## Notes

### Competing Interest Statement

The authors have declared no competing interest.

